# Persistence of psychotic experiences and clinical outcomes in adolescents at familial high risk of schizophrenia or bipolar disorder: The Danish High Risk and Resilience Study

**DOI:** 10.64898/2026.08.27.26361507

**Authors:** Sinnika Birkehøj Rohd, Anne Amalie Elgaard Thorup, Martin Wilms, Marta Schiavon, Doris Helena Bjarnadóttir Streymá, Andreas Færgemand Laursen, Anette Faurskov Bundgaard, Anne Søndergaard, Mette Falkenberg Krantz, Lotte Veddum, Carsten Hjorthøj, Aja Greve, Ole Mors, Merete Nordentoft, Nicoline Hemager, Maja Gregersen

## Abstract

**Objective:** This study examined the prevalence of psychotic experiences (PE) and how early onset and persistence of PE contribute to risk and severity of mental disorders in adolescents at familial high-risk of schizophrenia (FHR-SZ) or bipolar disorder (FHR-BP) and adolescents from a population-based control group (PBC).

**Methods:** This is the second follow-up of a nationwide cohort study including 522 children at FHR-SZ (N=202), FHR-BP (N=120), and PBC (N=200). Participants were assessed at ages 7, 11, and 15 using a semi-structured interview to evaluate PE and mental disorders.

**Results:** At age 15, adolescents at FHR-SZ reported more PE than PBC over the past six months (current) and the past four years, while adolescents at FHR-BP only reported more current PE. PE reported at two or three timepoints (persistent PE) predicted any Axis I disorder in mid-adolescence, corresponding to three- (OR 2.9, 95% CI [1.5-5.7]) and 21-fold (OR 21.4, 95% CI [2.8-162.3]) increased risks, respectively. Persistent PE also predicted multimorbidity, with three- (OR 2.8, 95% CI [1.0-7.6]) and four-fold (OR 4.1, 95% CI [1.2-14.1]) increased risks, respectively. This was after adjustment for sex, early mental disorders, and familial risk.

**Conclusions:** This study demonstrates a strong link between persistent PE and mid-adolescence mental disorders. Our findings emphasize PE as important risk markers for mental disorders during mid-adolescence and highlight the importance of monitoring children with PE before age 7 who develop persistent symptoms.

## 1. Introduction

While psychotic experiences (PE), encompassing hallucinations and delusions without a diagnosable psychotic disorder, are common in children and adolescents [1], persistent PE (i.e., PE present across repeated assessments) may represent a distinct developmental trajectory associated with mental disorders [2–4]. This question is particularly relevant in children at familial high-risk of schizophrenia (FHR-SZ) or bipolar disorder (FHR-BP), who are already at increased risk for mental disorders [5, 6]; the co-occurrence of familial liability and persistent PE may identify those at the highest risk for psychiatric outcomes.

PE are associated with increased risk of psychotic and non-psychotic disorders [7, 8], higher severity of non-psychotic mental disorders [9–11], poorer global functioning [12], lower quality of life [13], and suicidal ideation, suicide attempts, and self-harm [14, 15]. Compared with transient PE, individuals with persistent PE are at increased risk of mental disorders, transition to clinical psychosis, self-harm, and suicide attempts [2, 16–19]. While studies suggest that persistent PE are stronger predictors of later psychopathology, others highlight that transient PE may also have some significant predictive value [12, 20]. PE become more strongly associated with other psychopathology with advancing age, especially when they appear during adolescence or later [9]. However, very few studies have assessed PE prior to age 9 [21–23]. As a result, little is known on the significance of PE before middle childhood for the development of psychopathology in adolescence.

Knowledge on the predictive value of PE in children and adolescents at FHR-SZ and FHR-BP is limited as most studies are based on general population samples [3]. Research indicates that PE are more prevalent among children and adolescents whose parents have schizophrenia than among offspring without a parental history of the disorder [21, 23, 24]. By contrast, children and adolescents of parents with bipolar disorder do not appear to exhibit a similarly elevated prevalence of PE [21, 23, 25]. In the baseline and first follow-up of the Danish High Risk and Resilience Study (the current study), we found that children who reported PE in early childhood were at higher risk of having a mental disorder [23], and in middle childhood persistent PE were associated with increased odds of middle childhood mental disorders after accounting for familial risk [21]. Further, persistent PE predicted mental disorders non-differentially across children at FHR-SZ, FHR-BP and children from a population-based control group (PBC) [21].

To the best of our knowledge, this is the first study to examine early-onset and persistent PE in relation to adolescent mental health in a familial high-risk sample.

### 1.1. Aims

We aimed to investigate 1) the prevalence of PE from age 11-15 in adolescents at FHR-SZ or FHR-BP, and PBC, 2) associations between the level of persistence of PE from early childhood to mid-adolescence and the presence and multimorbidity of mental disorders in mid-adolescence, and 3) potential differential associations between the level of persistent PE and the presence and multimorbidity of mental disorders across adolescents at FHR-SZ or FHR-BP, and PBC.

## 2. Method

### 2.1 Participants

This study is the second follow-up of the Danish High Risk and Resilience Study (the VIA study), a longitudinal population-based cohort study of 522 seven-year-old children at FHR-SZ (*N*=202), FHR-BP (*N*=120) and PBC (*N*=200). Baseline assessments performed at age 7 were repeated with 4-year intervals, at ages 11 and 15. At the first follow-up (the VIA 11 study), 465 11-year-old children (FHR-SZ, *N*=179, FHR-BP, *N*=105, PBC, *N*=181) were re-assessed, and in this second follow-up (the VIA 15 study), a total of 427 15-year-old adolescents (FHR-SZ, *N*=158, FHR-BP, *N*=100, PBC, *N*=169) were re-assessed. The Danish Civil Registration System [26] and the Danish Psychiatric Central Research Register [27] were used to identify parents with or without a diagnosis of schizophrenia spectrum psychosis or bipolar disorder. The children at FHR-SZ had at least one parent with a schizophrenia spectrum psychosis: ICD-10 codes F20, F22 and F25, or ICD-8 codes 295, 297, 298.29, 298.39, 298.89 and 298.99. The children at FHR-BP had one or two parents with bipolar disorder: ICD-10 codes F30 and F31, or ICD-8 codes 296.19 and 296.39. Parents to PBC were not registered with either disorder. At baseline, PBC were matched with children at FHR-SZ on municipality, sex, and age. Children at FHR-BP were a non-matched group but comparable to the other two groups in terms of age and sex. Information on participants’ ethnicity was not collected. However, cohort inclusion required that both parents were born in Denmark to facilitate linkage to the national registers. The design of the VIA study is described in detail elsewhere [28–30].

The VIA 15 study was approved by the Danish Data Protection Agency (P-2019-273) and the Danish Committee on Health Research Ethics (H-20067908). The Danish Ministry of Health gave permission to retrieve data from Danish registers. Written informed consent was obtained from all adult participants and from the legal guardians of the participating adolescents. Assent was obtained from the adolescents.

### 2.2 Measures

#### 2.2.1. Assessment of psychotic experiences

The assessment of PE was conducted using the psychosis supplement of the semi-structured interview, the Kiddie Schedule for Affective Disorders and Schizophrenia – Present and Lifetime Version (K-SADS-PL) at all three timepoints [31]. Interviews were performed by trained psychologists, research nurses, and medical doctors, who were blinded to FHR-status. A primary caregiver was selected for each child, preferably the adult who spent the most time with the child and where the child’s address was registered. The child and primary caregiver were interviewed about nine types of hallucinations and 13 types of delusions. If the child’s or the caregiver’s answer received a score of 2 (possible psychotic symptom) or 3 (definite psychotic symptom), they were further interviewed about when the symptom last occurred, frequency, duration, impact on daily functioning, distress, and degree of conviction. All possible PE were rated on a seven-point scale (0=absent, 1=possibly present, 2=mild, 3=moderate, 4=moderately severe, 5=severe but not psychotic, 6=severe and psychotic) to ensure exclusion of uncertain symptoms. Ad modum Gregersen et al. [21], these scores were recoded into two categories: 1) absent or possibly present based on a score from 0 −1 or 2) definite PE based on a score from 2-6. We only included definite PE in this study. PE were consensus-rated at a clinical conference by a professor of child and adolescent psychiatry (second author, AAET) at the first and second follow-up. Hypnagogic and hypnopompic symptoms and symptoms attributed to fever or drugs were systematically excluded at the first and second follow-up. The definition of PE was slightly broader at baseline. The baseline study and the first follow-up are described in detail elsewhere [21, 23].

The interviews of PE are covering the children’s life from ages 0-15: baseline interviews addressed symptoms from age 0-7, the first follow-up covered symptoms from age 7-11, and the second follow-up covered symptoms from age 11-15.

From the questions regarding hallucinations, five variables were created: Auditory, loud thoughts, visual, olfactory, and tactile hallucinations. From the questions regarding delusions, 13 variables were created: Grandiose, guilt/sin, control, somatic, nihilism, thought broadcasting, thought insertion, thought withdrawal, messages from TV/radio, persecution, mind reading, reference, and other. Further, “any PE” was defined as the presence of any type of hallucination and/or delusion rated ≥2. Any PE from age 11-15 was divided into “current PE”, i.e., PE within the last 6 months, and PE within the past four years.

For the analysis of PE persistence, a categorical predictor variable was established, with four categories: children not reporting any lifetime PE (“never”), children reporting PE at one timepoint (“once”), children reporting PE at two timepoints (“twice”), and children reporting PE at all timepoints (“thrice”).

#### 2.2.2. Assessment of mental disorders

DSM-IV and V Axis I mental disorders were evaluated using the K-SADS-PL [31]. Assessments were conducted at all three time points, covering the children’s lives from birth until age 15. Interviews were conducted separately with the primary caregiver and the child, and the interviewers were blinded to FHR-status. The included mental disorders are detailed in Table 1 [6] [32] [33]. All diagnoses were confirmed with a specialist in child and adolescent psychiatry (second author, AAET).

**Table 1:**
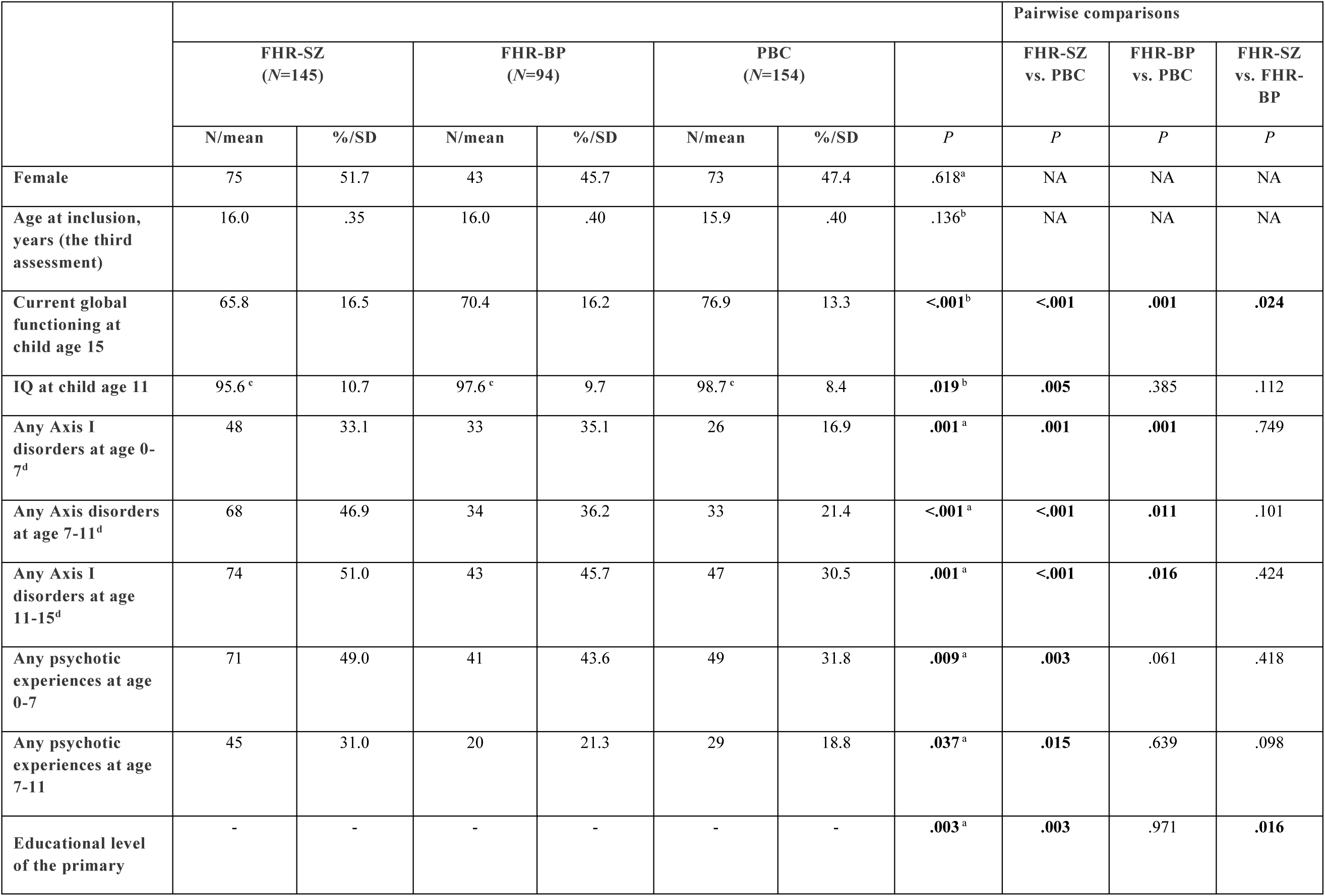
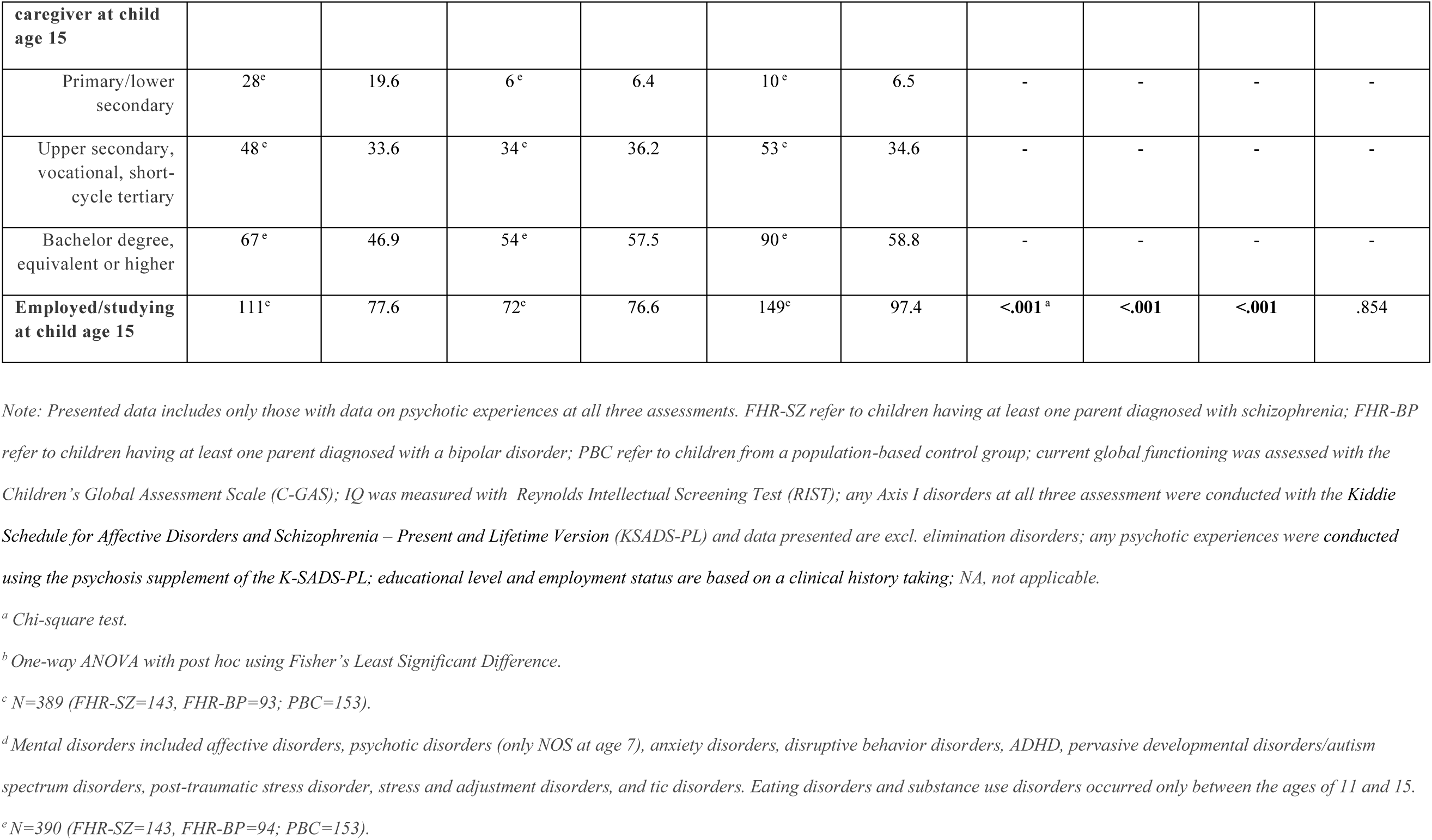
Sample characteristics of 393 adolescents and their primary caregivers in families with parental schizophrenia or bipolar disorder and a population-based control group.

Variables were created to indicate the presence of an Axis I mental disorder between age 0-7, 7-11, and 11-15. In addition, a variable indicating whether an Axis I mental disorder was present across age 0-11 was created. Multimorbidity was defined as presence of more than one mental disorder from age 11-15, i.e., one, or ≥two.

#### 2.2.3. Assessment of global functioning and general intelligence

The Children’s Global Assessment Scale (CGAS) [34] was used to evaluate the adolescent’s global functioning defined as psychological and social functioning over the previous month.

Scores range between 0 and 100, where lower values indicate poorer daily functioning. The Reynold’s Intellectual Screening Test (RIST) was used to assess children’s general intelligence at age 11 [35]. An index score reflecting an IQ estimate is derived from age-stratified normative data [36] with higher scores indicating better cognitive ability.

### 2.3. Statistical analysis

We compared demographic and clinical background characteristics of the three groups using chi-square tests for categorical data and one-way ANOVA for continuous data. To assess dropout, we conducted chi-square tests and independent samples t-tests.

To address our first aim, investigating the prevalence of PE from age 11-15, frequencies and percentages of PE were calculated using cross-tabulations. Group differences were analyzed using binary logistic regression, which was subsequently adjusted for sex. These analyses were conducted separately for the 4-year prevalence (age 11-15) and the 6-month prevalence. For our second and third aim, we applied logistic regression to examine the associations between level of PE persistence (i.e. PE once, PE twice, and PE thrice) and mid-adolescence Axis I mental disorders, using those who never experienced PE as the reference group. The analysis was subsequently adjusted for sex and early (age 0-7) and/or middle childhood (age 7-11) Axis I mental disorders. Potential interaction effects of FHR-status were examined by adding FHR to the adjusted models and if the interaction was non-significant, we also adjusted for FHR-status in the model. We did not adjust for socioeconomic status due to its close association with FHR-status. A binary logistic regression was conducted to examine whether PE reported in early childhood (age 0-7) predicted the presence of an Axis I mental disorder in mid-adolescence (age 11-15). We adjusted for sex, early childhood Axis I mental disorder and FHR-status (if the interaction with FHR-status was non-significant). A logistic regression analysis was conducted to assess whether the level of PE persistence was associated with multimorbidity at age 15, applying the same adjustment and interaction model as described above. To assess a potential dose-response relationship between persistence of PE and mid-adolescence Axis I mental disorders as well as multimorbidity, we examined linear trend by entering the PE persistence variable as an ordinal continuous predictor in a logistic regression model. Analyses were adjusted with the same adjustment and interaction model as described above.

The PE variables included children who received a psychosis diagnosis at some point across the three assessments. Sensitivity analyses were conducted excluding these cases.

Standard errors were clustered at the family level to account for sibling dependence in all models.

Statistical significance was defined as *p*<.05. Analyses were conducted using SPSS Statistics, version 29.0.1.0 (171) [37] and Stata/SE version 18.5 [38].

## 3. Results

### 3.1. Sample characteristics

The three groups were comparable with respect to sex and age at inclusion in the VIA 15 study. Adolescents at FHR-SZ or FHR-BP demonstrated significantly lower levels of global functioning compared with PBC, and the FHR-SZ group showed lower global functioning compared with FHR-BP. Children at FHR-SZ had a lower IQ at age 11 and more PE at age 7 and 11 compared with PBC, whereas both FHR-SZ and FHR-BP offspring were more likely to fulfill criteria for any Axis I disorder across all three assessments than PBC (Table 1). The study population included 414 adolescents in the prevalence analysis from age 11 to 15 and 393 children/adolescents in the analyses linking PE trajectories to psychopathology. A loss to follow-up analysis demonstrated that the 414 adolescents included in the prevalence analysis exhibited a higher level of global functioning than those who dropped out from the baseline study (*N*=108; t(512)=2.3, CGAS mean difference=3.9, 95% CI [.6-7.2], *P*=.020). There was no significant difference regarding prevalence of PE at age 7 (X^2^ (1)=1.7, *P*=.189, or regarding FHR group (X^2^ (2)=2.6, *P*=.270). A drop-out analysis comparing children who did not participate in all three assessments (*N*=129) with the 393 children included in the PE trajectory analysis showed that the latter had a higher level of global functioning at age 7 (t(512)=2.0, CGAS mean difference=3.1, 95% CI [.0-6.2], *P*=.048), and that there were no differences regarding prevalence of PE (X^2^ (1)=.1, *P*=.765), or FHR group (X^2^ (2)=2.2, *P*=.325).

### 3.2. Prevalence of PE in adolescence

A significantly higher proportion of adolescents at FHR-SZ reported any PE between age 11-15 compared with PBC. Adolescents at FHR-BP did not differ significantly from PBC or FHR-SZ. A higher proportion of adolescents at FHR-SZ or FHR-BP reported current PE compared with PBC, whereas FHR-SZ and FHR-BP did not differ significantly from each other (Table 2). Both hallucinations and delusions were more prevalent in adolescents at FHR-SZ compared with PBC from age 11 to 15 and over the past six months. Adolescents at FHR-BP reported significantly more delusions over the past six months compared with PBC (Table ST1).

**Table 2:**
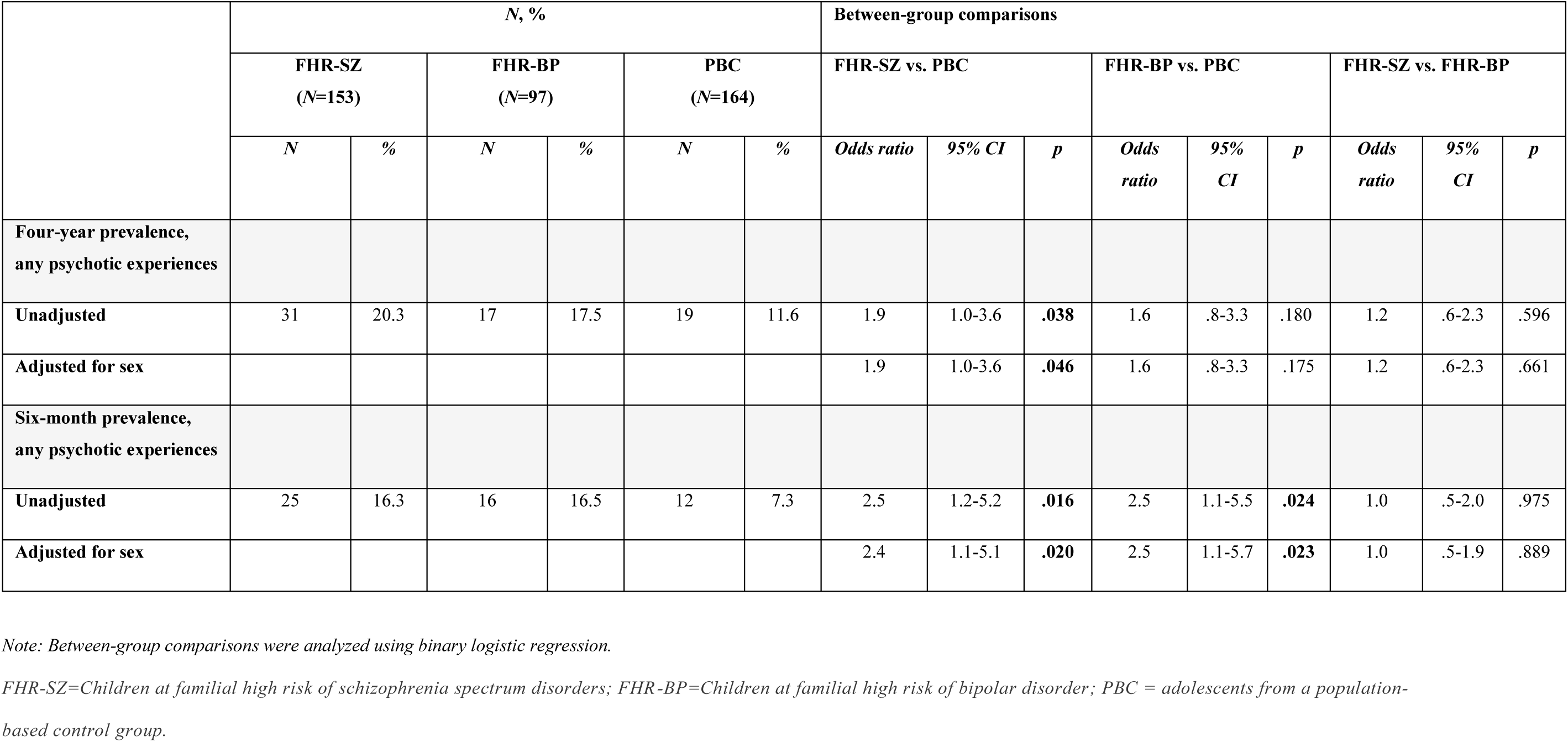
Psychotic experiences in adolescence (age 11-15) in 414 adolescents at familial high-risk for schizophrenia or bipolar disorder and adolescents from a population-based control group.

|  | *N*, % |  |  |  |  |  | Between-group comparisons |  |  |  |  |  |  |  |  |
| --- | --- | --- | --- | --- | --- | --- | --- | --- | --- | --- | --- | --- | --- | --- | --- |
|  | FHR-SZ<br>(*N*=153) |  | FHR-BP<br>(*N*=97) |  | PBC<br>(*N*=164) |  | FHR-SZ vs. PBC |  |  | FHR-BP vs. PBC |  |  | FHR-SZ vs. FHR-BP |  |  |
|  | *N* | % | *N* | % | *N* | % | *Odds ratio* | *95% CI* | *p* | *Odds ratio* | *95% CI* | *p* | *Odds ratio* | *95% CI* | *p* |
| Four-year prevalence,<br>any psychotic experiences |  |  |  |  |  |  |  |  |  |  |  |  |  |  |  |
| Unadjusted | 31 | 20.3 | 17 | 17.5 | 19 | 11.6 | 1.9 | 1.0-3.6 | **.038** | 1.6 | .8-3.3 | .180 | 1.2 | .6-2.3 | .596 |
| Adjusted for sex |  |  |  |  |  |  | 1.9 | 1.0-3.6 | **.046** | 1.6 | .8-3.3 | .175 | 1.2 | .6-2.3 | .661 |
| Six-month prevalence,<br>any psychotic experiences |  |  |  |  |  |  |  |  |  |  |  |  |  |  |  |
| Unadjusted | 25 | 16.3 | 16 | 16.5 | 12 | 7.3 | 2.5 | 1.2-5.2 | **.016** | 2.5 | 1.1-5.5 | **.024** | 1.0 | .5-2.0 | .975 |
| Adjusted for sex |  |  |  |  |  |  | 2.4 | 1.1-5.1 | **.020** | 2.5 | 1.1-5.7 | **.023** | 1.0 | .5-1.9 | .889 |

*Note:* Between-group comparisons were analyzed using binary logistic regression.
*FHR-SZ=Children at familial high risk of schizophrenia spectrum disorders; FHR-BP=Children at familial high risk of bipolar disorder; PBC = adolescents from a population-based control group.*

For distribution of hallucination and delusion types and PE persistence across adolescents at FHR-SZ, FHR-BP, and PBC refer to Figures SF1, SF2 and SF3.

After excluding five children with psychotic disorders, the difference between FHR-SZ and PBC on PE prevalence from age 11-15 was no longer significant. The differences between FHR-SZ and PBC on the six-month prevalences remained significant (Table ST2).

### 3.3. PE as predictors of Axis I disorders in mid-adolescence

PE reported at two or three time points significantly predicted the presence of any Axis I mental disorder in mid-adolescence. This association remained significant after adjusting for sex, early and/or middle childhood Axis I mental disorders, and FHR-status. The strongest association was observed for “PE trice”. There were no interaction effects with FHR-status (Table 3). A linear trend showed that the prevalence of Axis I mental disorders increased significantly across the PE persistence continuum (Table 3). In the “PE never” group, 27.5% had at least one Axis I mental disorder, whereas the proportion was 44.2% in the “PE once” group, 59.6% in the “PE twice” group, and 95% in the “PE thrice” group.

**Table 3:** Associations between trajectories of PE from early childhood to adolescence and the presence of mental disorders in adolescence (age 11-15) and the association between the degree of PE persistency and increasing diagnostic burden (comorbidity) of Axis I disorders in adolescence.

|  | PE once<br>N=154 (39.2 %) |  |  | PE twice<br>N=52 (13.2 %) |  |  | PE thrice<br>N=20 (5.1 %) |  |  | Linear trend<br>N=393 |  |  |
| --- | --- | --- | --- | --- | --- | --- | --- | --- | --- | --- | --- | --- |
|  | *Odds ratio* | *95% CI* | *p* | *Odds ratio* | *95% CI* | *p* | *Odds ratio* | *95% CI* | *p* | *Odds ratio* | *95% CI* | *p* |
| **Any Axis I disorder in adolescence<sup>a</sup>** |  |  |  |  |  |  |  |  |  |  |  |  |
| Unadjusted | 2.1 | 1.3-3.3 | **.002** | 3.9 | 2.0-7.5 | **<.001** | 50.0 | 6.5-387.2 | **<.001** | 2.3 | 1.8-3.0 | **<.001** |
| Adjusted for sex | 2.0 | 1.3-3.3 | **.003** | 3.8 | 2.0-7.2 | **<.001** | 44.2 | 5.6-349.5 | **<.001** | 2.3 | 1.8-2.9 | **<.001** |
| Adjusted for sex and early and/or middle childhood axis I disorder | 1.7 | 1.0-2.8 | **.043** | 3.0 | 1.5-6.0 | **.002** | 23.4 | 3.2-171.9 | **.002** | 1.9 | 1.5-2.5 | **<.001** |
| Adjusted for sex, early and/or middle childhood axis I disorder, and familial risk<sup>b</sup> | 1.6 | 1.0-2.6 | .074 | 2.9 | 1.5-5.7 | **.002** | 21.4 | 2.8-162.3 | **.003** | 1.9 | 1.4-2.5 | **<.001** |
| **Multimorbidity<sup>a</sup>** | N=68 (41.5 %) |  |  | N=31 (18.9 %) |  |  | N=19 (11.6 %) |  |  | N=164 |  |  |
| Unadjusted | 1.2 | .5-2.6 | .706 | 3.6 | 1.4-9.5 | **.009** | 5.0 | 1.5-16.1 | **.008** | 1.8 | 1.3-2.6 | **.001** |
| Adjusted for sex | 1.2 | .5-2.6 | .705 | 3.6 | 1.4-9.4 | **.009** | 4.9 | 1.5-16.1 | **.009** | 1.8 | 1.3-2.6 | **.001** |
| Adjusted for sex and early and/or middle childhood axis I disorder | 1.1 | .5-2.4 | .867 | 3.2 | 1.2-8.6 | **.020** | 3.8 | 1.1-12.9 | **.031** | 1.7 | 1.7-2.5 | **.006** |
| Adjusted for sex, early and/or middle childhood axis I disorder, and familial risk<sup>c</sup> | .9 | .4-2.1 | .824 | 2.8 | 1.0-7.6 | **.046** | 4.1 | 1.2-14.1 | **.023** | 1.7 | 1.2-2.5 | **.006** |

Note: Associations were analyzed using binary logistic regression with the reference group being PE never: N=167 (42.5 %) and 46 (28.1 %), respectively.
<sup>a</sup> Mental disorders included affective disorders, psychotic disorders, anxiety disorders, disruptive behavior disorders, ADHD, pervasive developmental disorders/autism spectrum disorders, post-traumatic stress disorder, stress and adjustment disorders, tic disorders, eating disorders and substance use disorders.
<sup>b</sup> No interaction with familial high-risk status: $P=.541$ and $P=.288$ for linear trend.
<sup>c</sup> No interaction with familial high-risk status: $P=.182$ and $P=.700$ for linear trend.

The sensitivity analysis removing the five children with psychotic disorder did not alter the results (Table ST3).

After adjusting for sex, Axis I mental disorder at age 7, and FHR-status, PE before age 7 predicted the presence of any Axis I disorder in mid-adolescence (1.5 OR, 95% CI [1.0-2.4], p=.0498), although the p-value was close to the conventional significance threshold. No interaction effect of FHR-status was observed (*P*=.256). To assess whether this association was driven by persistent PE, we conducted an exploratory logistic regression among children with PE at age 7. Those who subsequently ceased reporting PE at later assessments were compared with those who continued to report PE at age 11 and/or 15. Among children who reported PE at age 7, 40.4% continued to report PE at age 11 and/or 15. After adjusting for sex, Axis I mental disorder at age 7, and FHR-status, these children had 3.7-fold increased odds of having a mental disorder in mid-adolescence compared with children who did not report PE after age 7 (OR=3.7, 95% CI: 1.8-7.8, p=.001). No interaction effect of FHR-status was observed (*P*=.400). Of those who reported PE at two assessments (PE twice), 86.5% had first reported PE at age 7.

### 3.4. Associations between PE persistence and multimorbidity in mid-adolescence

A higher level of PE persistence was significantly associated with multimorbidity after adjusting for sex, early and/or middle childhood Axis I mental disorders, and FHR-status. A significant linear trend was observed across PE persistence levels and multimorbidity after adjusting for sex, early and/or middle childhood Axis I mental disorder, and FHR-status. There were no interaction effects with FHR-status (Table 3).

After removing four children with a psychotic disorder the association between PE twice and multimorbidity was rendered non-significant after all adjustments (Table ST3).

## 4. Discussion

### 4.1. Main findings

In this longitudinal cohort study, we found that current PE (within the last 6 months) and PE during the past four years were more often reported by adolescents at FHR-SZ compared with PBC. This third wave showed that adolescents at FHR-BP more often reported PE when measured within the last 6 months compared with PBC. After accounting for FHR-status, sex and childhood Axis I mental disorders, PE reported at two or three time points significantly predicted any Axis I mental disorder in mid-adolescence, with odds for having a mental disorder in mid-adolescence increasing threefold and twenty-onefold, respectively. Moreover, the odds increased with greater persistence of PE over time, suggesting a dose-response relationship. PE reported before age 7 as a predictor of mental disorders in mid-adolescence appeared to be largely driven by children whose PE persisted beyond age 7. Further, we found a dose-response relationship between persistent PE and multimorbidity, with an almost two-fold increase in odds for each level of PE persistence.

PE predicted all outcomes non-differentially across FHR groups.

### 4.2. Interpretation

The higher prevalence of PE observed in adolescents at FHR-SZ is in line with findings at baseline and first follow-up of the current cohort [21, 23], and with an independent cohort, suggesting robustness across samples [24]. The higher six-months prevalence in adolescents at FHR-BP has not previously been reported in this cohort and may suggest a delayed emergence of psychosis-related vulnerability in FHR-BP offspring. Future follow-ups will reveal the clinical significance of this finding. The finding that adolescents at FHR-SZ report elevated levels of both hallucinations and delusions, whereas adolescents at FHR-BP only report more delusions, may reflect underlying differences in vulnerability mechanisms, where adolescents at FHR-SZ show a broader neurodevelopmental disruption consistent with other findings from our cohort [39].

The associations between persistent PE and child psychopathology are consistent with previous literature [19, 21]. However, previous studies have not investigated the association between persistent PE reported at three time points and general psychopathology in mid-adolescence. The association between PE at age 7 and mental disorders in mid-adolescence appeared to be driven by children whose PE persisted into later childhood and/or adolescence, suggesting that persistence, rather than the early occurrence of PE per se, may be conferring the higher risk for mental disorders in mid-adolescence. Nevertheless, early assessments of PE before age 7 should be considered a relevant target group, as they provide the baseline from which persistence can be identified and examined. This is particularly relevant given that 86.5% of the children with moderately persistent PE (i.e., PE twice) reported PE at age 7. Had these children first been assessed at age 11, they would have been classified as having transient PE or none. Timely identification and support could potentially improve long-term outcomes.

A previous study has shown that the prevalence of PE increases in a dose-response pattern with the number of Axis I mental disorders [9]. Our study builds on this by taking the level of persistence into account, demonstrating a clear dose-response relationship, where multimorbidity increased with the number of time points of reported PE.

While adolescents at FHR-SZ or FHR-BP exhibited a higher prevalence of current PE compared with PBC, the association between PE persistence and mid-adolescence mental disorders and multimorbidity did not differ across FHR groups. Thus, PE appears indicative of a comparable risk for later psychopathology regardless of FHR-status, in accordance with previous findings [21, 40].

### 4.3. Future perspectives

The finding that the prevalence and multimorbidity of mental disorders in mid-adolescence increased significantly across the PE persistence continuum starting in early childhood can inform evidence-based decision-making in a clinical setting. The fact that 95% of those reporting PE at all three time points had an Axis I mental disorder in mid-adolescence highlights the clinical importance of identifying and monitoring PE in child development. At present, there is a lack of evidence-based prevention and intervention strategies based on identification of PE that can effectively reduce not only the risk of psychosis, but also broader psychopathology and its severity [3].

### 4.4. Strengths and limitations

This study has several strengths. We used a gold-standard interview to assess PE and Axis I mental disorders. To our knowledge, this is the first study to investigate PE at such an early age and to track their level of persistence and significance for mid-adolescence psychopathology, while also considering the role of FHR-status. Another strength is the narrow age range of the participants. Among the limitations is the smaller sample size of the FHR-BP group, which weakens the precision of estimates for this group. Additionally, we were not able to distinguish between parental bipolar disorder with and without psychosis, or to account for whether PE were experienced as distressing or non-distressing, which may have provided further insights into the prevalence and significance of PE in children and adolescents with varying degrees of psychosis risk.

### 4.5. Conclusions

Current PE and PE reported within the past four years were more frequently reported among adolescents at FHR-SZ compared with PBC, while adolescents at FHR-BP more often reported current PE than PBC. Future follow-ups will reveal the clinical significance of these findings.

This study demonstrates that PE reported prior to age 7 can serve as an early intervention target and provide a baseline from which persistent PE can be examined. Furthermore, children who continue to experience PE over time are highly likely to have a mental disorder in mid-adolescence and to present with multimorbidity, and these associations were observed non-differentially across FHR-status. These results suggest an opportunity for early identification of children at elevated risk for mental disorders which can facilitate timely intervention in both FHR groups as well as in the general population.

## Data Availability

Due to GDPR regulations, individual-level data cannot be shared publicly. However, researchers may apply for and receive approval to collaborate on the project.

## Acknowledgements

The authors gratefully acknowledge all the families who participated in the Danish High Risk and Resilience study. Further, we sincerely thank J. Ohland, L. Carmichael, M. Nymand, M. Enevoldsen, L. J. Mikkelsen, J. M. Brandt, Å. K. Prøsch, M. Birk, A. K. Andreassen, H. B. Stadsgaard, C. B. Knudsen, N. L. Steffensen, and A. M. Bundsgaard for contributing to the data collection, and C. B. Pedersen and M. G. Pedersen for assistance with data extraction from the Danish Registers.

This work was supported by the Lundbeck Foundation (R277-2018-594), Innovation Fund Denmark, Capital Region of Denmark, Mental Health Services of the Capital Region of Denmark, Aarhus University Hospital – Psychiatry, the Independent Research Fund Denmark (2096-00065B), Tryg Foundation, and Novo Nordisk Foundation (NNF20OC0060468).

