## Supplementary for "Persistence of psychotic experiences and clinical outcomes in adolescents at familial high risk of schizophrenia or bipolar disorder: The Danish High Risk and Resilience Study"

**Table ST1:** Hallucinations and delusions in adolescence (age 11-15) in children at familial high-risk for schizophrenia or bipolar disorder and children from a population-based control group.

|  | ***N*, %** | | | | | | **Between-group comparisons** | | | | | | | | |
| --- | --- | --- | --- | --- | --- | --- | --- | --- | --- | --- | --- | --- | --- | --- | --- |
|  | **FHR-SZ  (*N*=153)** | | **FHR-BP  (*N*=97)** | | **PBC  (*N*=164)** | | **FHR-SZ vs. PBC** | | | **FHR-BP vs. PBC** | | | **FHR-SZ vs. FHR-BP** | | |
|  | ***N*** | ***%*** | ***N*** | ***%*** | ***N*** | ***%*** | ***Odds ratio*** | ***95% CI*** | ***p*** | ***Odds ratio*** | ***95% CI*** | ***p*** | ***Odds ratio*** | ***95% CI*** | ***p*** |
| **Four-year prevalence, hallucinations** |  |  |  |  |  |  |  |  |  |  |  |  |  |  |  |
| **Unadjusted** | 26 | 17.0 | 11 | 11.3 | 15 | 8.5 | 2.2 | 1.1-4.4 | **.027** | 1.4 | .6-3.2 | .459 | 1.6 | .7-3.4 | .229 |
| **Adjusted for sex** |  |  |  |  |  |  | 2.1 | 1.1-4.3 | **.033** | 1.4 | .3-.9 | .455 | 1.6 | .7-3.4 | .262 |
| **Six-month prevalence, hallucinations** |  |  |  |  |  |  |  |  |  |  |  |  |  |  |  |
| **Unadjusted** | 19 | 12.4 | 7 | 7.2 | 9 | 5.5 | 2.4 | 1.1-5.7 | **.037** | 1.3 | .5-3.7 | .576 | 1.8 | .7-4.6 | .201 |
| **Adjusted for sex** |  |  |  |  |  |  | 2.4 | 1.0-5.6 | **.043** | 1.3 | .5-3.7 | .575 | 1.8 | .7-4.5 | .223 |
| **Four-year prevalence, delusions** |  |  |  |  |  |  |  |  |  |  |  |  |  |  |  |
| **Unadjusted** | 22 | 14.4 | 12 | 12.4 | 10 | 6.1 | 2.6 | 1.2-5.7 | **.019** | 2.2 | .9-5.2 | .083 | 1.2 | .6-2.5 | .655 |
| **Adjusted for sex** |  |  |  |  |  |  | 2.5 | 1.1-5.6 | **.023** | 2.2 | .9-5.3 | .080 | 1.2 | .5-2.5 | .722 |
| **Six-month prevalence, delusions** |  |  |  |  |  |  |  |  |  |  |  |  |  |  |  |
| **Unadjusted** | 17 | 11.1 | 11 | 11.3 | 7 | 4.3 | 2.8 | 1.1-7.1 | **.029** | 2.9 | 1.1-7.7 | **.036** | 1.0 | .4-2.2 | .956 |
| **Adjusted for sex** |  |  |  |  |  |  | 2.7 | 1.1-6.9 | **.034** | 2.9 | 1.1-7.7 | **.035** | .9 | .4-2.2 | .901 |

*Note: Between-group comparisons were analyzed using binary logistic regression.
FHR-SZ=Children at familial high risk of schizophrenia spectrum disorders; FHR-BP=Children at familial high risk of bipolar disorder; PBC = children from a population-based control group.*

**Figure SF1:** Prevalence of different types of hallucinations during adolescence across children at familial high-risk for schizophrenia or bipolar disorder and children from a population-based control group.

|  | Procent |
| --- | --- |
| FHR-SZ | 12.4 |
| FHR-BP | 5.6 |
| PBC | 3.7 |
| FHR-SZ | 2.6 |
| FHR-BP | 1 |
| PBC | 0 |
| FHR-SZ | 6.5 |
| FHR-BP | 2.1 |
| PBC | 1.8 |
| FHR-SZ | 3.9 |
| FHR-BP | 4.1 |
| PBC | 1.8 |
| FHR-SZ | 1.3 |
| FHR-BP | 1 |
| PBC | 1.2 |

Note: Bar chart.

**Figure SF2:** Prevalence of different types of hallucinations during adolescence across children at familial high-risk for schizophrenia or bipolar disorder and children from a population-based control group.

|  | Procent |
| --- | --- |
| FHR-SZ | 2 |
| FHR-BP | 1 |
| PBC | 0 |
| FHR-SZ | 1.3 |
| FHR-BP | 0 |
| PBC | 0 |
| FHR-SZ | 2 |
| FHR-BP | 0 |
| PBC | 0 |
| FHR-SZ | 0.7 |
| FHR-BP | 2.1 |
| PBC | 0 |
| FHR-SZ | 0 |
| FHR-BP | 0 |
| PBC | 0 |
| FHR-SZ | 1.3 |
| FHR-BP | 0 |
| PBC | 0 |
| FHR-SZ | 0 |
| FHR-BP | 3.1 |
| PBC | 0 |
| FHR-SZ | 0 |
| FHR-BP | 0 |
| PBC | 0 |
| FHR-SZ | 0 |
| FHR-BP | 1 |
| PBC | 0 |
| FHR-SZ | 9.8 |
| FHR-BP | 5.2 |
| PBC | 3.1 |
| FHR-SZ | 0.7 |
| FHR-BP | 0 |
| PBC | 1.2 |
| FHR-SZ | 0.7 |
| FHR-BP | 2.1 |
| PBC | 0.6 |
| FHR-SZ | 3.9 |
| FHR-BP | 1 |
| PBC | 1.2 |

Note: Bar chart.

**Figure SF3:** Distribution of psychotic experience (PE) persistency across adolescents at familial high-risk for schizophrenia or bipolar disorder and adolescents from a population-based control group.

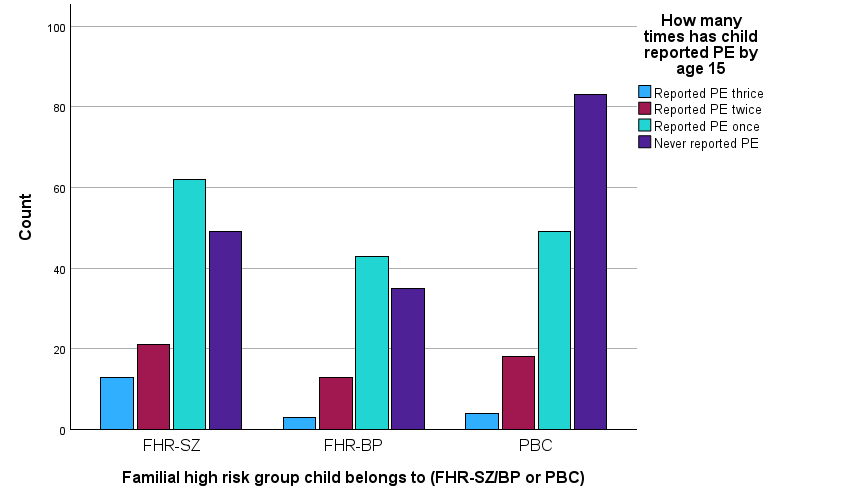

Note: Bar chart.

**Tabel ST2:** Psychotic experiences in adolescence (age 11-15) in children at familial high-risk for schizophrenia or bipolar disorder and children from a population-based control group. Excluding those with a lifetime psychotic disorder.

|  | ***N*, %** | | | | | | **Between-group comparisons** | | | | | | | | |
| --- | --- | --- | --- | --- | --- | --- | --- | --- | --- | --- | --- | --- | --- | --- | --- |
|  | **FHR-SZ  (*N*=149)** | | **FHR-BP  (*N*=96)** | | **PBC  (*N*=164)** | | **FHR-SZ vs. PBC** | | | **FHR-BP vs. PBC** | | | **FHR-SZ vs. FHR-BP** | | |
|  | ***N*** | ***%*** | ***N*** | ***%*** | ***N*** | ***%*** | ***Odds ratio*** | ***95% CI*** | ***p*** | ***Odds ratio*** | ***95% CI*** | ***p*** | ***Odds ratio*** | ***95% CI*** | ***p*** |
| **Four-year prevalence, any psychotic experiences** | 29 | 19.5 | 16 | 16.7 | 19 | 11.6 |  |  |  |  |  |  |  |  |  |
| **Unadjusted** |  |  |  |  |  |  | 1.8 | 1.0-3.4 | .055 | 1.5 | .7-3.1 | .247 | 1.2 | .6-2.4 | .581 |
| **Adjusted for sex** |  |  |  |  |  |  | 1.8 | 1.0-3.4 | .067 | 1.5 | .8-3.2 | .239 | 1.2 | .6-2.3 | .652 |
| **Six-month prevalence, any psychotic experiences** | 23 | 15.4 | 15 | 15.6 | 12 | 7.3 |  |  |  |  |  |  |  |  |  |
| **Unadjusted** |  |  |  |  |  |  | 2.3 | 1.1-4.8 | **.026** | 2.3 | 1.0-5.2 | **.038** | 1.0 | .5-2.0 | .968 |
| **Adjusted for sex** |  |  |  |  |  |  | 2.3 | 1.1-4.7 | **.032** | 2.4 | 1.1-5.4 | **.036** | .9 | .5-1.9 | .874 |

*Note: Between-group comparisons were analyzed using binary logistic regression.
FHR-SZ=Children at familial high risk of schizophrenia spectrum disorders; FHR-BP=Children at familial high risk of bipolar disorder; PBC = children from a population-based control group.*

**Table ST3:** Associations between trajectories of PE from early childhood to adolescence and the presence of mental disorders in adolescence (age 11-15), as well as the association between the degree of PE persistency and increasing diagnostic burden (comorbidity) of Axis I disorders in adolescence. Excluding those with a lifetime psychotic disorder.

|  | **PE once  *N*=152 (39.2 %)** | | | **PE twice *N*=50 (12.9 %)** | | | **PE trice *N*=19 (4.9 %)** | | | **Linear trend *N*=388** | | |
| --- | --- | --- | --- | --- | --- | --- | --- | --- | --- | --- | --- | --- |
|  | ***Odds ratio*** | ***95% CI*** | ***p*** | ***Odds ratio*** | ***95% CI*** | ***p*** | ***Odds ratio*** | ***95% CI*** | ***p*** | ***Odds ratio*** | ***95% CI*** | ***p*** |
| **Any axis I disorder in adolescence** |  | | |  | | |  | | |  | | |
| Unadjusted | 2.0 | 1.3-3.2 | **.003** | 3.9 | 2.0-7.7 | **<.001** | 47.3 | 6.1-365.9 | **<.001** | 2.3 | 1.8-3.0 | **<.001** |
| Adjusted for sex | 2.0 | 1.2-3.2 | **.004** | 3.9 | 2.0-7.5 | **<.001** | 40.8 | 5.2-321.4 | **<.001** | 2.3 | 1.7-2.9 | **<.001** |
| Adjusted for sex and early and/or middle childhood axis I disorder | 1.6 | 1.0-2.6 | .071 | 3.2 | 1.6-6.3 | **.001** | 21.3 | 2.9-155.5 | **.003** | 1.9 | 1.5-2.6 | **<.001** |
| Adjusted for sex, early and/or middle childhood axis I disorder, and familial risk^a^ | 1.5 | .9-2.5 | .112 | 3.1 | 1.5-6.1 | **.001** | 19.6 | 2.6-147.6 | **.004** | 1.9 | 1.4-2.5 | **<.001** |
| **Comorbidity** | ***N*=66 (41.3 %)** | | | ***N*=30 (18.8 %)** | | | ***N*=18 (11.3 %)** | | | ***N*=160** | | |
| Unadjusted | 1.1 | .5-2.4 | .877 | 3.4 | 1.3-9.0 | **.012** | 4.6 | 1.4-14.7 | **.011** | 1.8 | 1.2-2.6 | **.002** |
| Adjusted for sex | 1.1 | .5-2.4 | .876 | 3.4 | 1.3-9.0 | **.013** | 4.5 | 1.4-14.7 | **.013** | 1.8 | 1.2-2.6 | **.002** |
| Adjusted for sex and early and/or middle childhood axis I disorder | .9 | .4-2.2 | .897 | 3.0 | 1.1-8.2 | **.030** | 3.3 | 1.0-11.0 | .052 | 1.6 | 1.1-2.4 | **.010** |
| Adjusted for sex, early and/or middle childhood axis I disorder, and familial risk^b^ | .8 | .3-1.9 | .621 | 2.6 | .9-7.2 | .068 | 3.6 | 1.1-12.1 | **.037** | 1.7 | 1.1-2.4 | **.010** |

*Note: Associations were analyzed using binary logistic regression with the reference group being PE never: N=167 (43.0 %) and N=46 (28.8 %), respectively.
^a^ No interaction with familial high-risk status: P=.558 and P=.305 for linear trend.*

*^b^ No interaction with familial high-risk status: P=.161 and P=.614 for linear trend.*
